# MYPP trial: Myo-inositol supplementation to prevent pregnancy complications in women with polycystic ovary syndrome, study protocol for a multicenter, double-blind, randomized controlled trial

**DOI:** 10.1101/2025.04.01.25325030

**Authors:** Anne W.T. van der Wel, Chryselle M.C. Frank, Rebekka Bout-Rebel, Ruben G. Duijnhoven, Bo E. van Bree, Olivier Valkenburg, Salwan Al-Nasiry, Robbert H.F. van Oppenraaij, Tatjana E. Vogelvang, Michelle E.M.H Westerhuis, Hedwig P. van de Nieuwenhof, Susanne C.J.P. Gielen, Myrthe L. Bandell, Mireille N. Bekker, Maurice G.A.J. Wouters, Velja Mijatovic, Arie Franx, Cornelis B. Lambalk, Frank J.M. Broekmans, Sabina de Weerd, Jet M.H. Gerards, Jelle H. Baalman, Jeroen van Disseldorp, Josje Langenveld, Marlise N. Gunning, Marinus J. Eijkemans (deceased), Geert W.J. Frederix, Rebecca C. Painter, Bart C.J.M. Fauser, Joop S.E. Laven, Bas B. van Rijn

## Abstract

**Introduction:** Pregnant women with polycystic ovary syndrome (PCOS) have an increased risk of gestational diabetes mellitus (GDM), preeclampsia and preterm birth, and their offspring are more likely to have an abnormal birth weight and require hospitalization. Current preventive strategies to reduce the rate of pregnancy complications associated with PCOS have ineffective. Myo-inositol supplementation has shown promising clinical benefit in preventing adverse outcomes in individuals with obesity and PCOS-related disorders. Recent randomized controlled trials using daily supplementation with 4 grams of myo-inositol during pregnancy estimated a relative reduction of up to 65% for GDM and preterm delivery.(1–7) These studies included both obese patients and patients with normal body weight. From these trials, post-hoc analysis suggested similar, or even greater, reductions in the sub group of women with PCOS.(8) However, no randomized controlled trials have been conducted to evaluate the potential benefit of myo-inositol supplementation in preventing pregnancy complications in these women.

**Methods and analysis:** This is a prospective, multicenter, double-blind, randomized controlled trial to study the clinical effectiveness and cost-effectiveness of myo-inositol supplementation to prevent pregnancy among women with PCOS. Recruitment will set out to include 464 individuals with PCOS between 8+0 and 16+0 weeks of pregnancy, who are then randomized in a 1:1 ratio to either the intervention group that will receive 4 grams myo-inositol added a standard recommended dose of folic acid, divided over two daily sachets of sugary powder throughout pregnancy; or into the control group that will receive similar looking sachets containing no supplement other than the routinely recommended dose of folic acid. Regular multivitamin use (without inositols) will be allowed for both groups. The primary endpoint is the incidence of the composite outcome of either GDM, preeclampsia and/or preterm birth.

Secondary endpoints include indicators of maternal physical and mental well-being, maternal health-related quality of life, neonatal outcomes, breastfeeding practices and breastmilk composition. Trial results will be analysed according to the intention-to-treat principle. A full economic evaluation will be performed.

**Ethics and dissemination:** The study protocol has been approved by the Medical Ethics Committee of the Erasmus MC University Medical Centre Rotterdam. Approval by the boards of management for all participating hospitals will be obtained. Trial results will be submitted to peer-reviewed journals.

**Trial registration number:** NTR-NL7799

**Strengths and limitations of this study:**

- This will be the first prospective randomized controlled trial, aimed specifically at women with polycystic ovary syndrome (PCOS), to estimate the potential of myo-inositol supplementation in preventing common pregnancy complications associated with PCOS.
- The double-blinded randomized multicenter design enables unbiased interpretation and generalisability of results.
- This study will provide novel recommendations on myo-inositol supplementation as a safe nutritional intervention aimed to improve pregnancy outcomes among women with PCOS.
- Inclusion of individuals of diverse ethnic backgrounds may prove a limitation given the relative homogeneous population of the Netherlands; the estimated effect sizes, compared with previous studies upon which our power calculations were based, may also vary across sub populations e.g. women with obesity and different PCOS phenotypes.
- Recruitment and compliance in this patient group may lead to self-selection of participants with higher educational, or socio-economic backgrounds. This needs to be taken into account to estimate the impact of implementation.

## INTRODUCTION

Polycystic ovary syndrome (PCOS) is the most common endocrine disorder in women of reproductive age with a reported prevalence of 6 - 15%, depending on the diagnostic criteria used.(9, 10) PCOS is a heterogeneous condition characterized by the combination of ovulatory dysfunction, either clinical or biochemical signs of hyperandrogenism, and/or the presence of polycystic ovary morphology on ultrasonography.(10) Apart from its reproductive consequences, which are primarily due to ovulatory dysfunction, PCOS can negatively affect psychological well-being, mainly through increased anxiety and depression, and is strongly associated with obesity and cardiometabolic changes including impaired glucose tolerance, type-2 diabetes mellitus, hypertension and the metabolic syndrome.(11–14). Insulin resistance and compensatory hyperinsulinemia, which affects both obese (70 – 80%) and lean (20-25%) women with PCOS, is a key feature of PCOS, and can further promote excess ovarian androgen production. Combined, these changes in metabolic health negatively affect periconceptional health as well as long-term healthy aging.(13, 15, 16) Multiple meta-analysis have consistently demonstrated an increased risk of pregnancy complications in people with PCOS, including gestational diabetes mellitus (GDM; odds ratio (OR) 3.7), pregnancy-induced hypertension (PIH; OR 5.5), preeclampsia (OR 3.5), and preterm birth (PTB; OR 1.8).(17, 18) As a consequence, children born to women with PCOS have a higher risk of an abnormal birth weight, admission to the neonatal intensive care unit (NICU: OR 2.3), and perinatal death (OR 3.1).(17, 19) Mechanisms to explain the link between PCOS and adverse pregnancy outcomes are not well understood, although key contributors appear to be periconceptional hyperandrogenism, increased fasting insulin and high glucose levels that are all associated with pregnancy complications.(20–22) Further, PCOS is thought to have a negative impact on breastfeeding success and breastmilk composition, although it is not clear from previous studies how much of this risk is explained by the coexistence of obesity.(23–26) Similar, long-term consequences of PCOS during pregnancy on offspring health and development are likely to occur, although this is yet an understudied area of research. Intrauterine exposure to even mild hyperglycemia is known to be associated with an increased risk of childhood adiposity, diabetes, and a greater likelihood of developing cardiovascular disease later in life.(27–29) In a limited number of studies, children of women with PCOS were indeed shown to have an increased risk of cardiovascular, endocrine, and respiratory disorders (30–32). Taken together, these results suggest that interventions to reduce the impact of PCOS on pregnancy outcomes have the potential to improve both perinatal health, as well as long term health outcomes beyond later in life.(30–33) Previous strategies to improve pregnancy outcomes among women with PCOS have not been very successful in reducing the incidence of major pregnancy complications. These have included several lifestyle intervention strategies, both with emphasis on dietary modification as well as stimulation of physical activity, that failed to show sustained positive effects on markers of metabolic disturbance, on body mass index (BMI), on biochemical hyperandrogenism, and on glucose and lipid levels.(34) Similar, a number of pharmacological interventions have unsuccessfully tried to improve pregnancy outcomes in women with PCOS, in particular with insulin-sensitizing agents e.g. metformin.(35, 36) However, none of these studies to date have been effective in reducing pregnancy complicationsm and safety concerns exist of such broad pharmacotherapeutic approach in pregnancy due to uncertainty about the risk of teratology, unwanted effects on fetal development, tolerability etc.(37)

In the past few years, evidence has emerged to support a role for myo-inositol supplementation as a safe nutritional intervention to reduce the risk of adverse pregnancy outcomes related to metabolic health. Inositols are naturally occurring substances present in many foods, with a structure similar to glucose. Inosistols are known to support several physiologic processes in virtually all living organisms and to act as second messengers in multiple biological processes. Myo-inositol, one of the nine stereo-isomeric forms of inositol, is often described as a natural insulin-sensitizer as it fulfils a second messenger role in the insulin signalling pathway by enhancing the translocation of GLUT-4 receptors on the plasma membrane and thereby facilitating the intracellular uptake of glucose.(38) This nutrient, historically considered part of the vitamin-B complex and consider a sugar alcohol, can be derived from many food products, fruits, and vegetables in particular, and can safely be used as a dietary supplement.(39) Technically, myo-inositol is considered a nonessential nutrient, because it is also synthesised *de novo* from glucose-6-phosphate in many tissues. Under normal circumstances, about 4 grams of myo-inositol is produced daily by the kidney alone. However, endogenous synthesis is not sufficient to maintain its wide use in metabolic processes, and an adequate intake of food is essential for its availability.(40) For example, an intake of about 100 mg of caffeine (the equivalent of about 2 cups of espresso coffee) can already lead to a temporary shortage of myo-inositol and requires a compensatory intake of inositol-rich foods (or supplements).

Evidence from previous studies suggests a rationale for optimizing myo-inositol levels, either by altering food intake or by supplementation in a number of conditions, including PCOS and obesity.(41, 42) In women with PCOS, signs of intracellular myo-inositol deficiency, as well as increased urinary excretion of myo-inositol, has been demonstrated, further supporting the rationale for supplementation.(43) In seven randomized controlled trials (RCTs), summarised by Zeng and colleagues, myo-inositol supplementation compared with placebo (or with metformin) in women with PCOS was associated with a decrease in indices of insulin resistance, e.g. homeostatic model assessment (HOMA) score(44), decreased levels of insulin, as well as decreased levels of luteinizing hormone and testosterone, signs of improved ovarian function, and improvements in cardiovascular risk profile e.g. lower blood pressure, lower lipids and reduced body-mass index.(45–47)

In pregnancy, low myo-inositol levels are associated with signs of increased insulin resistance (i.e. type-2 diabetes mellitus), higher blood glucose levels in GDM and preeclampsia.(48–50) To date, seven RCTs have been performed using myo-inositol supplementation from the first trimester to prevent pregnancy complications.(1–7). Four of these trials were conducted in a Southern European population, among women at risk of GDM (N=828) who were either overweight, obese or had a family history positive for type-2 diabetes mellitus. In these trials, participants received supplements containing 4 grams of myo-inositol per day, added to the standard recommended dose of 0.4 mg folic acid, 12-13 weeks of gestation onwards.(1, 2, 6, 7). The use of myo-inositol was associated with 58-60% lower rates of GDM as well as a reduction in preterm birth, macrosomia and infants born large-for-gestational-age (LGA). In another RCT by Farren and colleagues, conducted in a single tertiary hospital in Ireland among women with a family history of diabetes, a combination of the two isoforms myo-inositol and D-chiro-inositol (containing a lower dose of myo-inositol of about 1.1 grams per day) was not associated with lower GDM rates (4). In all seven studies myo-inositol supplementation was well tolerated and no side effects were reported, except a single study reported complaints of headache after use of myo-inositol.(3) No RCTs have been published to evaluate the effectiveness of myo-inositol supplementation to prevent adverse pregnancy outcomes in women with PCOS, which is the subject of this study.

### Aim

The primary objective of this study is to assess whether daily supplementation with myo-inositol during pregnancy among women with PCOS can reduce the composite outcome GDM, PTB, and preeclampsia.

The secondary objectives of the trial are to evaluate the effect of myo-inositol on the incidence of other adverse pregnancy outcomes (e.g. indicators of maternal and neonatal health, birth weight, and biomarkers of insulin and glucose homeostasis, endocrine factors, mental health, and other relevant patient-reported outcomes), as well as cost-effectiveness analysis and budget-impact analysis of myo-inositol supplementation as a preventative strategy for pregnant women with PCOS.

Based on the previously reported beneficial effects observed in (non-pregnant) women with PCOS, as well as the positive results observed in preventing pregnancy complications in other study populations, we hypothesize that myo-inositol supplementation is a safe and promising intervention to be tested in a robust clinical prevention trial.

## METHODS AND ANALYSIS

The flow of the trial is shown in *Figure I* - Study timeline and procedures (Standard Protocol Items: Recommendations for Interventional Trials (SPIRIT) Figure.

### Trial design and setting

The MYPP-trial is designed as a prospective, multicenter, double-blind, randomized controlled intervention trial among pregnant women with PCOS. The trial will be coordinated from the Erasmus MC University Medical Center Rotterdam (sponsor) and will be conducted at 13 study sites, including 3 university medical centres and 10 non-academic teaching hospitals in the Netherlands. An updated list of participating centres can be found on the study website (www.mypp-trial.nl).

### Patient and public involvement

The Dutch PCOS Foundation (Stichting PCOS) and members of the Dutch Patient Organisation for Couples with Fertility Problems (Freya) were involved in the design of the MYPP-trial and have agreed to remain actively involved during trial conduct, e.g. by creating awareness for the study, participating in the trial project group meetings and reviewing information provided to patients. Similar feedback was acquired through a focus group consisting of individual PCOS patients. Finally, a preliminary survey was conducted among patients with PCOS to determine whether the proposed study was feasible and accepted.

### Study population

#### Participants and eligibility criteria

Patients are eligible to participate when they have been diagnosed with PCOS according to the Rotterdam consensus criteria, and the following 2018 International evidence-based guideline for the assessment and management of PCOS (criteria are provided in Table I);(10, 30): a singleton pregnancy; confirmed viability (positive heartbeat on ultrasound); and pregnancy between 8+0 and 16+0 weeks gestational age. Additional inclusion criteria are maternal age ≥18 years, sufficient command of the Dutch or English language and willingness to provide written informed consent.

**Table I.** PCOS diagnostic criteria.

|  | Definition and cutoff-value |
| --- | --- |
| 1. Oligo-/amenorrhoea | ▪ *Oligomenorrhoea*: 35-182 days<br>or<br>▪ *Amenorrhoea*: > 182 days |
| 2. Hyperandrogenism | ▪ *Clinical*: hirsutism defined as Modified Ferriman Gallwey score $\geq 4$-6 (whether or not accompanied by acne and alopecia)<br>and/or<br>▪ *Biochemical*: elevated Calculated Free Testosterone or Free Androgen Index > 4.5 (FAI = (total testosterone x 100)/SHBG) |
| 3. Polycystic ovaries on transvaginal sonography | ▪ *Volume*: one or two ovaries > 10cm<sup>3</sup><br>and/or<br>▪ *Follicle count* (2-9mm): one or two ovaries $\geq 12$ follicles per ovary<sup>[2]</sup> |

AND
B. *Other aetiologies of the above criteria are excluded*, such as thyroid disease, hyperprolactinemia, androgen-secreting tumors, Cushing's syndrome and non-classing congenital adrenal hyperplasia\*.
\* Exclusion of other causes requires TSH, Prolactin levels, FSH and, if clinical status indicates other causes need to be excluded.

Women with pre-existing type 1 or type 2 diabetes mellitus, renal failure (as defined by a glomerular filtration rate of below 50 mL/min), or other serious chronic kidney disease or systemic disease that is thought to interfere with myo-inositol metabolism or excretion will be excluded. Women using myo-inositol supplements (and wished not to discontinue these supplements) are excluded, as well as women using other insulin-mimetics, hypoglycaemic agents, and systemic steroids.

### Recruitment

In the case of fertility treatment for PCOS (estimated to be ∼80% of patients with PCOS), information on the study is provided before conception and informed consent procedure was organised after confirmation of a viable pregnancy. In addition, patients may be enrolled at any of their antenatal clinical appointments between 8+0 and 16+0 weeks gestational age (either after spontaneous conception or after fertility treatment). Written informed consent is obtained by Good Clinical Practice (GCP) trained research midwives, physicians or investigators.

### Intervention

Women in the intervention arm of the trial will receive supplementation with 4 grams myo-inositol added to the standard recommended dose of 0.4 mg folic acid, divided over two daily doses. Women in the control arm will receive standard supplementation of 0.4 mg folic acid daily only. The dietary supplements are formulated as a slightly sweet-tasting finely granulated powder in 2 daily sachets, with resembling sensory characteristics for both groups. Supplements were custom-made and provided by the manufacturer (Gedeon Richter, Belgium) following instructions specified for the trial and Good Medicinal Practice-certified safety and manufacturing protocols. The sachets of powder may be added to food, diluted in a glass of water or juice, or consumed directly. All participating women will receive sufficient instructions, from trained research staff, on the proper use of the supplementation during their pregnancy. In countries of the European Union, myo-inositol (EC-number 201-281-2) and folic acid (EC-number 200-419-0) are both registered as dietary (food) supplements and authorized to be used as agents to optimize a healthy diet. Folic acid supplements are part of the standard-of-care recommendation for the purpose of reducing the risk of having a pregnancy affected with spina bifida or other neural tube defects and have previously been used as a complementary therapy in trials on myo-inositol supplementation in pregnancy and PCOS patients.(51, 52)

Myo-inositol is affirmed as Generally Recognized as Safe (GRAS) as a direct human food ingredient.(39) The quantities of myo-inositol supplementation proposed are considered standard amounts, comparable to quantities synthesised de novo in healthy human and consistent with the dosage used in previous trials in which myo-inositol supplementation was shown to be safe and effective without any side-effects reported. Dosage modifications are not required or performed.

Myo-inositol supplementation in the context of this clinical trial is not considered as a clinical trial of an investigational medicinal product, or CTIMP, by the Dutch and EU regulations, and this protocol has been confirmed as such by Central Committee on Research Involving Human Subject (CCMO) governing body of the Netherlands.

### Randomisation, blinding and allocation

After obtaining informed consent, local investigators will randomly allocate participants to either the intervention group or the control group by providing the appropriate supplements. For this, the indistinguishable boxes of supplements will be labelled with sequential randomisation numbers according to the randomisation list (allocation sequence) generated using the online software tool ALEA (accessible at https://nl/tenalea/net). Block randomisation will be performed on a 1:1 basis, with permuted blocks of the random block size (sizes 4 and 6). To prevent any imbalance between groups in aspects of care that may differ between participating sites, randomisation will be stratified by site. To ensure that the study is blinded to trial participants, care providers, and researchers, labelling of the study supplements and randomisation is carried out by an independent research staff member and statistician at the Erasmus Medical Centre Rotterdam, both not involved in further research or patient care. Until blinding is broken, the meaning of the randomisation numbers (i.e. information on treatment allocation) is solely accessible to the independent research staff member.

### Concomitant care and interventions

During the trial, routine obstetrical care and treatment of pregnancy complications following national guidelines will be permitted. As part of standard antenatal care and regardless of randomisation group, all pregnant women receive nutritional and lifestyle advice in agreement with Dutch guidelines.(53, 54) Participants may use regular over-the-counter products, such as multivitamins, except for those supplements containing myo-inositol.

### Outcomes measures Primary outcome

The primary outcome of our trial is the number of participants in both study arms that reach the composite endpoint of either i) gestational diabetes mellitus, and/or ii) preeclampsia and/or iii) preterm birth (i.e. birth before 37 weeks). The final outcome will be scored up to 1 week after delivery, to account for women who develop postpartum preeclampsia. GDM is defined according to Dutch national guidelines as any degree of glucose intolerance with onset or first recognition during pregnancy.(55) Diagnosis is made in accordance with local protocol in participating centres. In practice, GDM is most commonly diagnosed using a 75-gram oral glucose tolerance test (OGTT) with either the WHO (1999) criteria for diagnostic thresholds of venous plasma glucose values (fasting ≥7.0 mmol/L or 2-hour post load ≥7.8 mmol/L), or the International Association of Diabetes and Pregnancy Study (2013) criteria of fasting glucose ≥ 5.1 and <7.0 mmol/l, or 1-hour glucose ≥10.0 mmol/l. Both criteria were deemed acceptable for this study, as both are widely used and reflect current clinical practice, which is transitioning towards more evidence-based criteria for cost-effective care for GDM. In the post-hoc analysis, additional analysis to evaluate the relative contribution of the diagnostic criteria will be performed. Preeclampsia is defined according to the revised definition of the International Society for the Study of Hypertension in Pregnancy as new-onset hypertension >140 mmHg systolic or >90 mmHg diastolic blood pressure after 20 weeks gestational age and the coexistence of one or more of the following new onset conditions: i) proteinuria, and/or ii) other maternal organ dysfunction: renal insufficiency, liver involvement, liver involvement, neurological complications, haematological complications and/or iii) uteroplacental dysfunction: fetal growth restriction.(56)

### Secondary outcomes

Secondary maternal outcomes are gestational hypertension, the diagnosis of haemolysis, elevated liver enzymes, and low platelets (HELLP)-syndrome, maternal mental health and health-related quality of life up until six weeks postpartum. Maternal mental health and health-related quality of life will be assessed using the following validated questionnaires at study inclusion, 36 weeks of GA and 6 weeks postpartum: Beck Depression Inventory Scale II Netherlands (BDI-II-NL) questionnaire (57), the 5-Level EuroQol - five dimension (EQ-5D-5L) questionnaire (58) and the Polycystic Ovary Syndrome Health-Related Quality of Life Questionnaire (PCOSQ). (59, 60) Fetal growth and utero vascular development will be assessed using ultrasound scans, including uterine artery Doppler measurements. Additional secondary neonatal outcomes include the incidence of infants with large-for-gestational-age (LGA) and small-for-gestational-age (SGA) birth weight, congenital abnormalities, neonatal hypoglycaemia and neonatal medium care or intensive care admissions. In addition, a full cost-effectiveness analysis will be performed.

### Additional study parameters and follow-up

Levels of markers representative of glucose homeostasis, endocrine balance and lipid profile will be measured in maternal blood at multiple time points and in cord blood. Urine samples will be collected to analyse myo-inositol excretion. Additional data concerning maternal health, obstetric and neonatal outcomes, compliance and tolerability of the nutritional intervention, breastfeeding practices and breastmilk composition will be assessed during the trial. Samples will be collected of maternal blood, urine, breast milk, umbilical cord blood, umbilical cord and placental tissue for future use in ancillary studies, as part of the Erasmus MC biobank regulation and facility, for which optional additional informed consent will be obtained. Permission for follow-up of children until school-age to monitor the long-term effects on development, metabolic changes and epigenetic changes shall be obtained (subject to additional protocols and funding).

### Study procedures Participant timeline

The study timeline and procedures are also presented in Figure I.

**Figure I.**
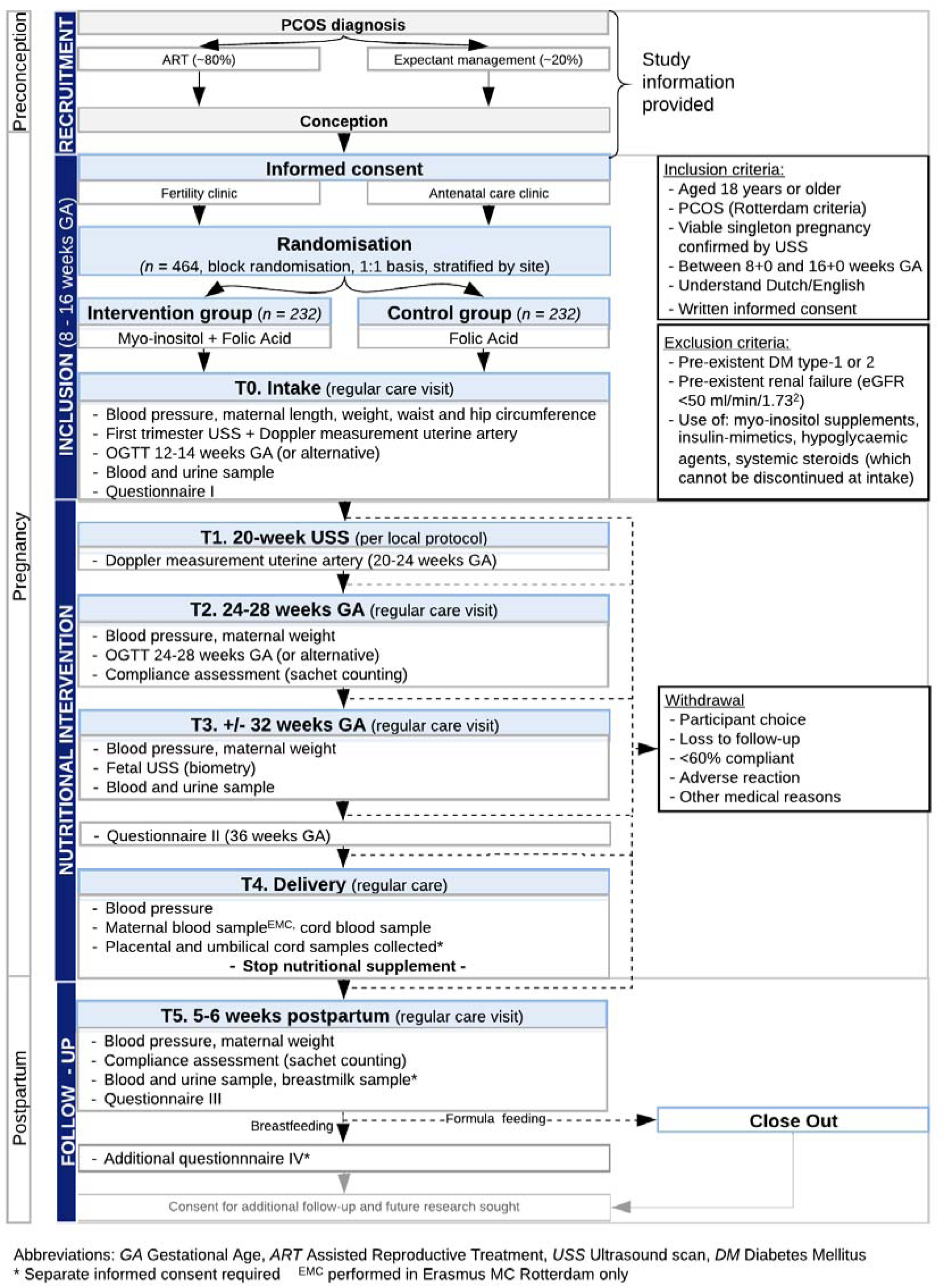
Study timeline and procedures (Standard Protocol Items: Recommendations for Interventional Trials (SPIRIT) Figure)

Study procedures will mostly coincide with routine antenatal care visits. After the confirmation of a viable pregnancy between 8+0 and 16+0 weeks by ultrasound, participants proceed through the process of informed consent and randomisation as described above. A local investigator will provide the allocated supplements and give instructions for appropriate use. During this first study visit maternal baseline characteristics and body anthropometry (blood pressure, maternal height and weight, hip and waist circumference) are ascertained and blood and urine samples are taken.

Following international guidelines on PCOS, we recommend assessing glycaemic status before 20 weeks gestation, preferably by using a 75-gram OGTT between 12 and 14 weeks gestation.(30) Any congenital abnormalities or polyhydramnios identified during a standard or advanced 20-week ultrasound scan will be recorded. A Doppler measurement of the uterine artery will be performed in the first trimester and repeated between 20 and 24 weeks gestational age. During the second study visit, between 24 and 28 weeks of gestation, blood pressure and maternal weight is recorded and compliance with the study supplements will be assessed by sachet counting. Between 24 and 28 weeks gestation, an OGTT is performed (per local protocol) to ascertain the primary outcome. Women who refuse OGTT testing or have a contra-indication will be offered an alternative (e.g. serial glucose monitoring). At the 32-week visit, anthropometry is repeated, and fetal growth scanning by ultrasound is performed. Participants are advised to discontinue the dietary supplements after labour and delivery. During the last study visit, around five to six weeks postpartum, the remaining supplements will be collected, maternal blood pressure and weight is measured and additional blood and urine samples will be taken.

### Research on breastfeeding

We will seek permission for follow-up of breastfeeding mothers until six months postpartum. After obtaining separate informed consent, breast milk samples will be collected five to six weeks postpartum for nutrient analysis and hormone and myo-inositol levels will be measured. Participants will receive a questionnaire six months postpartum to gather information on current (breast)feeding practices, maternal (mental) health and lifestyle and paternal baseline characteristics.

### Data collection and biological specimens

Data regarding outcomes, including maternal and neonatal morbidity and hospital admissions, will be collected up to six weeks postpartum (up to six months postpartum in case of participation in breastfeeding research). Participants will receive three study questionnaires, to be completed at study inclusion, 36 weeks of gestational age and 6 weeks postpartum. These study questionnaires will be a composite of different validated questionnaires to assess lifestyle (including physical activity (Short QUestionnaire to ASsess Health enhancing physical activity (SQUASH)(54)), maternal mental health (BDI-II-NL)(57), maternal health-related quality of life (EQ-5D-5L)(58) and PCOSQ(59, 60), compliance with the dietary supplements used for this study (Eight-Item Morisky Medication Adherence Scale (MMAS-8) (61), cost-effectiveness of the nutritional intervention (iMTA Medical Consumption Questionnaire (iMCQ)(62), iMTA Productivity Cost Questionnaire (iPCQ)(63). In addition, questions addressing demographic characteristics, nutritional status (including use of additional dietary supplements), PCOS diagnosis and associated symptoms, and breastfeeding intentions and practices were incorporated. Between 24 to 28 weeks of gestation and 6 weeks postpartum, compliance with the nutritional intervention will be assessed by sachet counting. In addition, information on compliance, patients’ overall satisfaction with supplementation and any issues encountered with consumption will be gathered by using a participant diary and via the study questionnaires at 36 weeks of gestation and six weeks postpartum.

Data collection will be done by trained research staff using an electronic case record form (eCRF) and collected in the access-controlled, web-based database OpenClinica. For participants who are unwilling or unable to comply with the protocol (including attendance at study visits and participants who discontinue or deviate from intervention), consent will be obtained to follow-up on key outcome measures.

### Safety and data monitoring

Considering the minimal risks associated with the use of myo-inositol supplements and in line with previous studies, the study is formally classified as of ‘negligible risk’ requiring ‘minimal monitoring’ following ICH guidelines for Good Clinical Practice (2005/28/EC). Nonetheless, as the study involves an intervention during pregnancy, the study will be additionally monitored for safety by an external independent Data and Safety Monitoring Board (DSMB). All serious adverse events (SAEs) occurring between randomization and six weeks postpartum will be reported to the accredited Medical Research Ethics Committee (MREC) and DSMB. An interim-analysis will be performed after 100 included participants have completed the study. This analysis will be executed by an independent statistician, unblinded for intervention allocation, who will report to the DSMB. Premature termination of the study will be decided if there is sufficient ground that continuation of the study will jeopardise the health or safety of the participants. The trial conduct will be audited by an external independent study monitor at each site, overseen by the study sponsor, following the minimal intensive risk-based monitoring plan established for this study.

### Sample size

The components of the primary composite outcome measure, a diagnosis of either GDM, PTB and/or pre-eclampsia are anticipated to occur in 23%, 4% and 9% of patients respectively, based on previous Dutch study cohort data.(21) Using this data, the incidence of the composite outcome can be estimated at 33% for our study population. Previous RCTs in pregnancy on myo-inositol supplementation at a dose of 4 grams, commenced in the first trimester, among women at risk of GDM (i.e. women with overweight, obesity or a family history of type-2 diabetes mellitus) reported a significant relative risk reduction of 58–60% in incident GDM. In a secondary analysis of these studies, a significant relative risk reduction of 55% was observed for preterm birth and a trend towards reduced risk for gestational hypertension (RRR 55%, p = 0.03). Trials were conducted in the Italian population with limited diversity. Due to the geographic distribution of citizens of non-Western origin in the recruitment area, we do expect to recruit a more diverse and multi-ethnic population. To avoid overestimation and take into account therapy compliance and differences in the study population, we aim for a more conservative goal of a 35% reduction in the composite outcome of GDM, preeclampsia, and preterm birth among pregnant women with PCOS. Based on a 35% relative risk reduction of the composite outcome and statistical power of 80% (α=0.05), the required sample size was calculated at n=464 (n=232 in each study arm).

### Analysis and reporting of results Primary and secondary outcomes

The primary analysis of the trial results will be performed according to the intention-to-treat principle. If a participant is unable to complete the entire course of the study as specified in the protocol, she will not be withdrawn from the study if consent is obtained to follow up on key outcome measures. The database will be validated by examining internal consistency and identifying data outliers. For missing data, the complete-case analysis will be used. Only in the event of high missing rates or in case missing’s are not at random, multiple imputation methods will be considered. Generally, *p*-value of <0.05 will be considered significant. Results will be reported according to Consolidated Standards of Reporting Trials (CONSORT) guidelines.

Secondarily, *a priori* sensitivity analysis will be performed, omitting participants who are withdrawn because of reluctance to continue with the trial or because they are unwilling or unable to take more than 60% of the interventional supplements, evidenced by sachet counting. When appropriate, and upon the advice of the trial statistician, further sensitivity analysis ‘per protocol’ or ‘as treated’ will be performed. The Statistical Analysis Plan (SAP) will be finalised by the project team of the study prior to database closure.

For the primary analysis, the number of individuals reaching the composite primary endpoint will be presented as exact numbers and percentages for each study arm and between-group analysis will be performed using the chi-square test (or Fishers Exact Test as appropriate). The effectiveness of the intervention will be presented as absolute risks, relative risks, relative risk reductions (along with 95% confidence intervals) and by assessing the number needed to treat (if applicable). Categorical secondary outcomes and additional study parameters will be compared between the two groups in the same way as the primary outcome. Continuous parameters will be presented as medians (with interquartile ranges) if data is skewed and as means (with standard deviations) if normally distributed. For these parameters, the independent *t*-test (if the outcome is normally distributed) or a non-parametric Mann-Whitney U-test (if the outcome is skewed) will be used to assess the potential differences between the two study arms. Predefined sub-group analysis will be performed comparing women with and without hyperandrogenism, as this is thought to be of relevance to the mechanism of action (and possibly efficacy) of myo-inositol supplementation.

### Economic evaluation

We forecast that the nutritional intervention will result in two categories of cost-savings: i) a decrease in costs for the treatment of pregnancy-related complications and neonatal morbidity/mortality, ii) a decrease in productivity impairment of mothers and/or fathers. The cost-effectiveness analysis (CEA) of the intervention will be performed according to the intention-to-treat principle. To evaluate cost-effectiveness from a societal perspective, all resources used in maternity and neonatal care from initiation of supplementation up until six weeks postpartum are recorded and multiplied by the cost per unit. The iMCQ will be used in the study questionnaire at 36 weeks gestational age and six weeks postpartum, as described previously, to include medical consumption outside the hospital.(62) To validate our cost analysis of pregnancy complications and long-term outcomes in the PCOS patients at baseline (control group), data from a recent study of our group (21) will be collected, in which the cost of PCOS-related complications and long-term outcomes were collect prospectively in at least 180 PCOS patients. To assess cost-effectiveness from a patient perspective, the iPCQ (63) and questions derived from the EQ-5D-5L (58) and the PCOSQ (59, 60) to assess maternal healthy-elated quality of life, are incorporated in the study questionnaire at study inclusion, 36 weeks of pregnancy and six weeks postpartum.

Both costs and effect measures will be combined in a so-called decision tree to calculate incremental cost-utility ratios (ICERs). Deterministic and probabilistic sensitivity analyses will be performed to demonstrate and visualize the uncertainty of our outcome measures. This will also result in the costs averted per complication and the total costs in both groups. Budget impact analysis of implementing the nutritional intervention into the Dutch healthcare system (i.e. recommending myo-inositol supplementation in addition to standard recommended folic acid supplementation) will be assessed in line with NZA guidelines through (decision analytical) modelling and analysed in a probabilistic way, addressing the following perspectives: i) societal perspective (using societal-CEA based prices), ii) health insurance and third party perspectives (using Dutch Healthcare Authority average rates and other applicable rates) and iii) health care perspective (using Ministry of Health, Welfare and Sport commissioned average rates).

## ETHICS AND DISSEMINATION

The trial protocol has been approved by the Medical Research Ethics Committee (MREC) of the Erasmus MC University Medical Centre, Rotterdam under the number MEC-2019-000. Approval by the Board of Directors will be obtained for all participating hospitals. The trial is registered in the public Dutch Trial Registry (NTR) under the number NTR-NL7799 in June 2019. MYPP is also registered online at ClinicalTrials.gov under the number NCT05524259 in September 2022. Substantial changes made to the research will be submitted for approval to the accredited MREC and updated on the NTR website. The full study protocol, including amendments, is publicly available on the study website (www.mypp-trial.nl). Data collection and handling will be done by GCP trained research staff in accordance with the General Data Protection Regulation (GDPR). To ensure patients’ privacy all data will be encoded (pseudononymised) using the unique non-speaking randomisation number that is randomly assigned in the process of treatment allocation. The encryption key of the randomisation code for linkage to personal information will only be available to the local research staff of the participating centres. Specimens taken for biobanking will be encoded and centrally stored at the biobank of the Erasmus MC in accordance with the code of conduct for responsible use (2011). After completion of the trial, the principal investigator will report on the results of the main analysis and submit a manuscript to a peer-reviewed journal. Further dissemination will occur through presentations at international meetings, and results will be shared through various communication channels in collaboration with the Dutch PCOS patient foundation.

## Data Availability

All data produced in the present study are available upon reasonable request to the authors

## Contributors

Study concept, trial design and study protocol: CMCF, RBR, MNG, GWJF, MJCE, TEV, CBL, FJMB, AF, BCJMF, JSEL, BBVR

Acquisition of data: CMCF, RBR, MLB, SDW, VM, MGAJW, MEMHW, JMHG, TEV, SSCJPG, RMFVDW, JPDB, HPVDN, RHFVO, JHB, SAN, BVB, OV, JVD, MNB, CBL, FJMB, AF, JSEL, BBVR

Analysis and interpretation of data: CMCF, RBR, GWJF, MJCE, JSEL, BBVR Drafting of the manuscript: CMCF, RBR, GWJF, MJCE, AF, JSEL, BBVR

Critical revision of the manuscript for important intellectual content: CMCF, RBR, MLB, JVD, SAN, RHFVO, RMFVDW, TEV, CBL, FJMB, AF, JSEL, BBVR

Study supervision: JSEL, BBVR

## Provenance and peer review

Not commissioned; externally peer reviewed.

## Open access

This is an open access article distributed in accordance with the Creative Commons Attribution Non Commercial (CC BY-NC 4.0) license, which permits others to distribute, remix, adapt, build upon this work non-commercially, and license their derivative works on different terms, provided the original work is properly cited, appropriate credit is given, any changes made indicated, and the use is non-commercial. See: http://creativecommons.org/licenses/by-nc/4.0/.

